# Making sense of publicly available data on COVID-19 in Ireland

**DOI:** 10.1101/2020.05.13.20101089

**Authors:** G. Mills, W. Cullen, N. Moore-Cherry, R. Foley

**Affiliations:** School of Geography, UCD, Dublin Ireland; School of Medicine, UCD, Dublin Ireland; Department of Geography, Maynooth University, Maynooth Ireland

## Abstract

This paper reports on the management of the COVID19 pandemic in Ireland over the period March to May 2020. During this period the Irish government implemented a series of policies designed to delay the spread of the pandemic culminating in a stay-at-home order that greatly restricted mobility for the majority of the population. In this paper we evaluate the policies enacted using evidence obtained from a number of novel sources, including census and real-time traffic data. The evidence suggests that the policies have impeded the spread of the virus, which has mostly been confined to Dublin and its commuter belt. At the same time, the virus has become concentrated in a number of clusters associated with nursing homes and workplaces that remained open during the delay phase. This evidence is used to hypothesise on the likely impact of the pandemic on high density and poor neighbourhoods in Dublin.

## Introduction

The COVID-19 outbreak was declared a global pandemic by the World Health Organisation on 11th March 2020. As of 30th April, over 3 million cases had been reported worldwide, and 228,000 people had died as a result of infection. On 29th February, the first positive case was reported in Ireland and by the end of April, there were nearly 20,000 reported cases and nearly 1000 deaths. In each country, management of the outbreak has been handled differently depending on the empirical evidence of its spread and modelling scenarios of the transmission within the population. The empirical evidence includes data on the rates of infection in the population and in sub-populations based on nature and place of work. The tools used everywhere include a mixture of testing, contact tracing and degrees of social isolation, including quarantining. The testing is done to find out who has the virus; the tracing is designed to identify who has been in contact with someone who has the virus and isolation is employed to separate out those infected or likely to be infected from the general population. Ideally, in the case of a global pandemic, where every country is affected, the empirical evidence would be gathered in a consistent manner so that the effectiveness of policies could be gauged and lessons learned. This has clearly not been the case for COVID where the empirical evidence is highly variable across countries making knowledge transfer difficult.

Internationally an important part of the public health response has been the engagement with the population through the exchange of information (including some data) on the strategies deployed and their effectiveness. This exchange has been essential to establishing public trust while enacting measures that impact on people’s livelihoods and social interactions. This paper examines relevant publicly available data for the six-week period (16/3/20 to 30/4/20) to describe the evolution of the COVID-19 pandemic in Ireland and the policies enacted to mitigate its impact.

Ireland has followed a strategy broadly in line with the World Health Organization and the European Centre for Prevention and Disease Control (ECDC), which has three components:

1. Containment Strategy: All efforts are focused on identifying cases (without regard to severity) and their contacts early, in order to prevent further transmission (secondary spread).
2. Delay Phase: This accepts the inevitability of transmission and is designed to slow down the spread such that the health system can cope with the consequences.
3. Mitigation Strategy: This is to be activated where containment is no longer effective in controlling the spread when the focus shifts to identifying the cases who are most severely unwell.

The health response to COVID-19 has been managed by the National Public Health Emergency Team (NPHET), which meets weekly to assess the international data, receive guidance regarding the outbreak and to review Ireland’s ongoing preparedness in line with advice from the WHO and the ECDC. The empirical evidence, including cases, hospitalisation and death rates, is assembled by the HSE Public Health and HSE Health Protection Surveillance Centre. The HSE High Consequences Infectious Diseases Planning and Coordination Group (HCID) has been working at a detailed level on this situation since early January and has put in place detailed plans and issued guidance and information in preparedness across the health service. An Expert Advisory Group was established in early February to provide advice to the National Public Health Emergency Team, the HSE and others on an ongoing basis.

## Methods

In Ireland, the first case of COVID-19 infection was confirmed on 29th February 2020. As of the 30^th^ April, the total number of confirmed cases nationally was 20,742 and 1050 people had died from the infection.

We undertook a secondary analysis of publicly available data reported in the six-week period from March 16th, when detailed information was first published to April 30th. The organisation of these data follow a standard pattern with slight deviations. On a daily basis, Ireland’s Department of Health publishes NPHET’s Statements based on data gathered up to 48 hours beforehand^1^. This includes summary information on: cases; hospital statistics, gender and age and, how the virus is spreading. In addition to this information, there are reports on: the Epidemiology of COVID-19 generated by the Health Protection Surveillance Centre (HPSC) and COVID-19 Daily Operations Update reports created by the Acute Hospitals Performance Management and Improvement Unit^2^. In the following we compile the data from these sources to provide an overview of the pandemic in Ireland and the policy responses. Table 1 shows a timeline of the pandemic and of the responses during the period of this study.

**Table 1.**
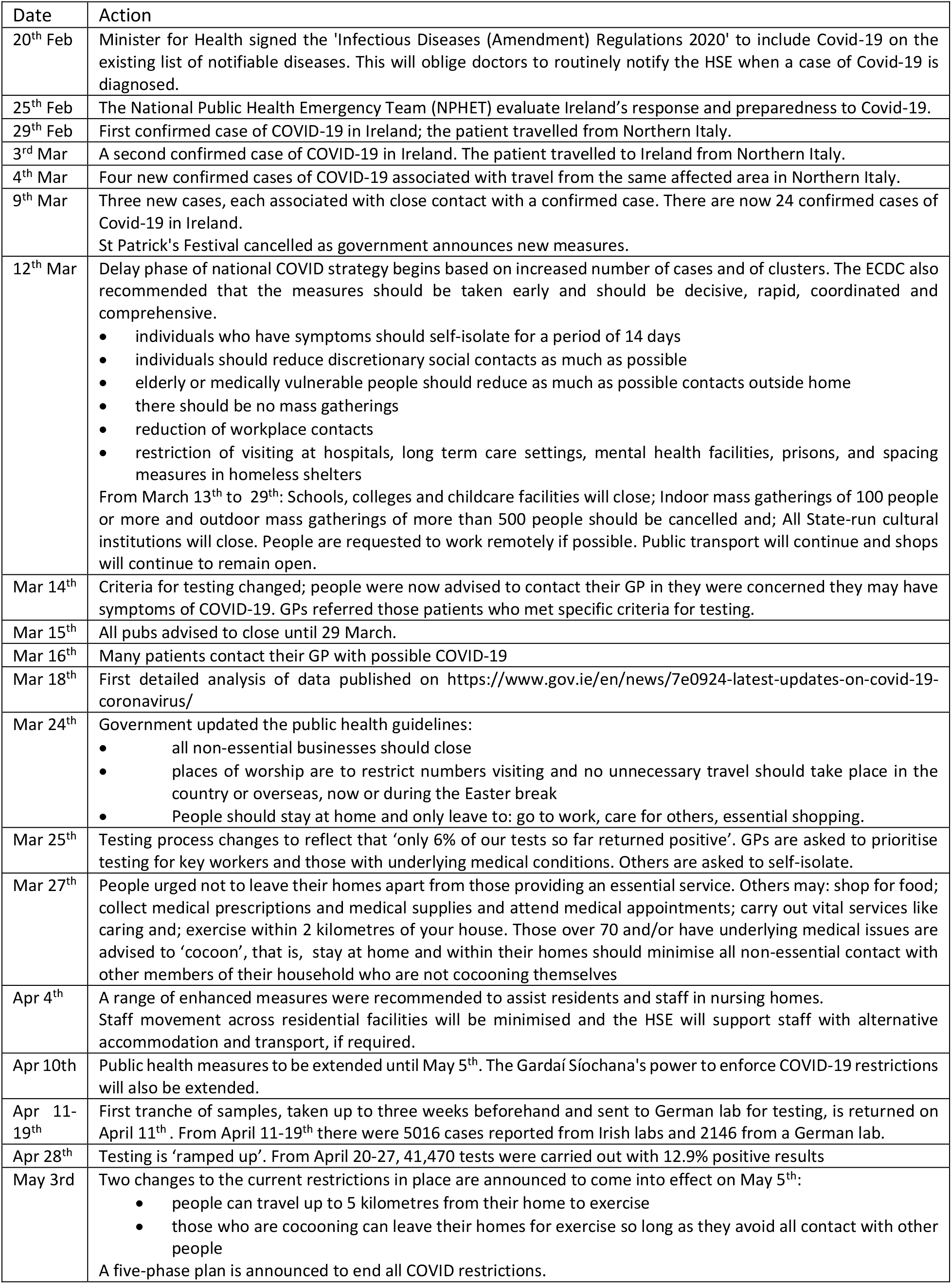
A timeline of responses to the COVID-19 pandemic in Ireland.

To interpret the temporal and spatial patterns in these data, we used available information from other sources. These include the 2016 Household Census^3^, which provides information on population, socio-economic characteristics and commuting patterns and real-time traffic information^4^. The population information from the census information is used to place the infection rate across the county into context and to examine the potential for spread within high-density areas. The traffic information is a very good measure of mobility, which can spread the virus.

## Results

We start Ireland’s COVID story on April 30^th^, two months after the first case was identified and then outline how it got to this point.

Table 2 reports the number of infections and hospitalisation by age group and compares the percentages in each category with the population as a whole. As is clear, there are infections among all age groups and the median age for cases is 49 years; in terms of gender, 58% of cases were female. However, the severity and outcomes are progressively worse with age. The death rate is concentrated in the oldest age group; as of 30^th^ April, over 90% of COVID-19 related deaths were aged 65+ years. These results are consistent with that reported in other countries.

**Table 2.** COVID19 cases, hospitalisations, admission to intensive care units (ICU) and deaths by age group as of April 30^th^, 2020. The final two columns indicates the population by age-group for Ireland.

| *Age*<br>(years) | *Cases* |  | *Hospitalised* |  | *ICU* |  | *Deaths* |  | *Census population* |  |
| --- | --- | --- | --- | --- | --- | --- | --- | --- | --- | --- |
|  | N | % | N | % | N | % | N | % | N (000s) | % |
| <5 | 101 | 0.5 | 16 | 0.6 | 0 | 0.0 | 0 | 0.0 | 315.3 | 6.4 |
| 5 – 14 | 205 | 1.0 | 8 | 0.3 | 1 | 0.3 | 0 | 0.0 | 693.7 | 14.1 |
| 15 – 24 | 1363 | 6.6 | 56 | 2.0 | 6 | 1.6 | 2 | 0.2 | 618.1 | 12.6 |
| 25 – 34 | 3271 | 15.9 | 173 | 6.3 | 13 | 3.5 | 4 | 0.4 | 620 | 12.6 |
| 35 – 44 | 3491 | 17.0 | 232 | 8.4 | 32 | 8.7 | 9 | 0.9 | 776.9 | 15.8 |
| 45 – 54 | 3752 | 18.3 | 368 | 13.3 | 79 | 21.5 | 20 | 1.9 | 661.6 | 13.4 |
| 55 – 64 | 2706 | 13.2 | 416 | 15.0 | 103 | 28.1 | 46 | 4.5 | 539.7 | 11.0 |
| 65+ | 5596 | 27.3 | 1496 | 54.0 | 133 | 36.2 | 952 | 92.2 | 696.3 | 14.1 |
| Total | 20510 |  | 2768 |  | 367 |  | 1033 |  | 4921.5 |  |

Of the 1,011 deaths that were confirmed as COVID-19 related, 45.8% were hospitalised, 5.6% were admitted to ICU; 86.8% had underlying medical conditions. 53% of these were male and 47% female and the median age was 81. Not all COVID cases are treated in the hospitals. There are 371 clusters and 4,590 cases in community residential settings, 219 (3679) of which were in nursing homes. 348 people were admitted to hospital from community residential settings, 233 of whom came from nursing homes. A total of 735 people have died in community residential settings (including probable deaths), 630 of whom were in nursing homes. This represents 59.7% and 51.1% of total COVID-19 related deaths in Ireland, respectively.^5^

As of April 28^th^ there were 5,627 cases in healthcare workers. Of these 3.7% were hospitalised and just 0.6% required treatment in ICU. There have been 5 deaths. The gender split for these cases is 27% male and 73% female. Of these infected workers, 34% were nurses, 24% were allied healthcare professionals, 24% were healthcare assistants, 7% were doctors and 1% were porters.

### Temporal trends

The headline figure presented on a daily basis is the number of new positive cases, which is taken as an indicator of the progression of the pandemic in Ireland. To put these values in context, it is important to consider the application of the testing system.

- Prior to March 13th, people with symptoms of acute respiratory infection and who had travelled to an area with confirmed COVID infections or who had been in contact with a confirmed case, were tested for COVID infection.
- After this date the criteria for testing changed such that people with acute respiratory symptoms were advised to contact their general practitioner who referred them for testing and this became standard practice from Monday 16th March.
- On March 25th, the guidelines were once again amended on the basis that hitherto, just 6% of the samples had tested positive. From this date samples were only offered to those in an at-risk group.

At risk groups included: close contact with a confirmed case; healthcare workers; persons with diabetes, immunosuppressed, chronic lung disease, chronic heart disease, cerebrovascular disease, chronic renal disease, chronic liver disease and smokers; staff or residents in a long-term care facility; prison staff and inmates where it may be difficult to implement self-isolation advice and; pregnant women. From March 25th, those not in the designated at-risk groups who contacted their GPs with symptoms that could be COVID related were advised to self-isolate for 14 days from the time of symptom onset, the last five of which should be symptom free. They also advised household contacts to restrict movements for 14 days after person has self-isolated.

Figure 1 shows the time series of cases and deaths nationally; by April 30^th^ the infection rate (that is, the number of reported cases divided by the population) was 43.56 per 10,000. Given the changes to the testing regime, we can expect that this rate is too low. This is particularly the case as it is possible to acquire and transmit the virus even though showing none of the common symptoms (such as high temperature, coughing and difficulty breathing). Because of such asymptomatic cases, the only means of getting a reliable estimate of the infection rate is to conduct random sampling of the entire population. As such, we can take the count as an indicator of the level of infection rather than a precise measure; however, the close temporal correspondence between the number of deaths and cases does show that these counts have some value in terms of indicating the severity of the cases.

**Figure 1.**
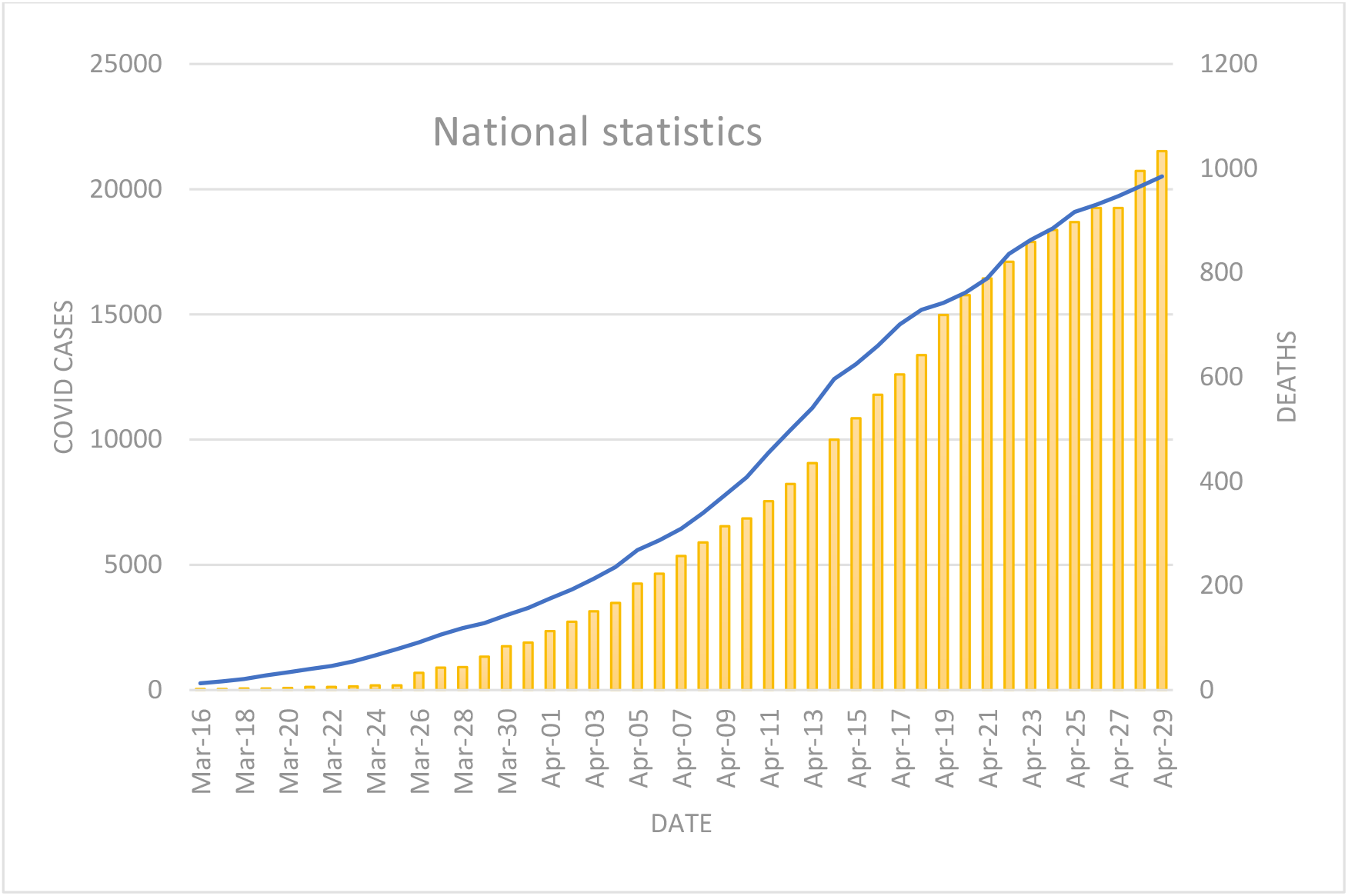
The number of reported positive COVID19 cases and related deaths accumulated since March 16^th^.

Figure 2 shows day-to-day pattern of new cases and reported deaths for the same period. Also shown is a 7-day moving average for the new cases. It is difficult to assess changes in the spread of the pandemic for a number of reasons, including the changing testing regime. For a period of several weeks in March and April, the capacity to process tests at Irish laboratories was exceeded and samples were sent to a German lab. These test results were added to the results from Irish labs between April 11–19^th^ adding 2146 cases to the totals, even though these dated from weeks beforehand. This would mean that the counts for March are lower (and those for April are higher) than the graphs depict.

**Figure 2.**
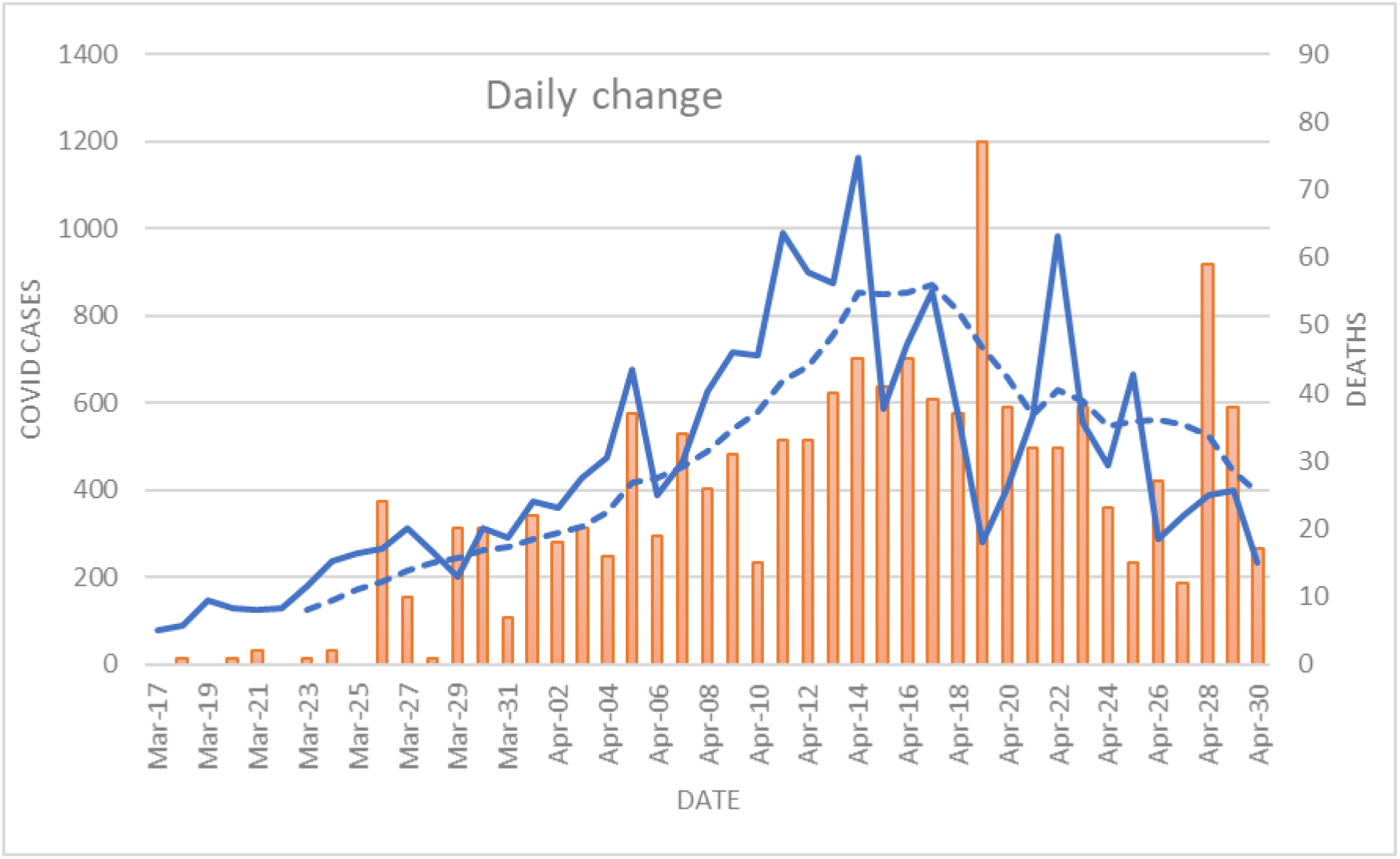
The daily change in COVID cases (blue line) and deaths; the broken blue line shows the 7 day moving average.

### Geographical trends

Case data is publicly available at the scale of the county. Figure 3 shows the number of cases by county for March 31^st^ and April 30^th^, corresponding to 3281 and 20,742 cases nationally, respectively. The labels on the map show infection rates (per 10,000) based on county population; on March 30^th^ the national rate was 7.0 per 10,000 and on April 30^th^ it was 44.3. The maps illustrate the rapid spread of reported cases across the county but also shows its concentration in Dublin and surrounding counties. To illustrate, the population of Ireland is 4.76 million and 28.3% (1.34 million) of these live in Dublin; by comparison, the second most populous county is Cork with 0.54 million (11.4%). So, if the infection had spread evenly, one would expect that about 11% of cases should be based in Cork but this is clearly not the case. On March 31^st^ there were 1838 (12.2 per 10,000) cases in Dublin but just 272 (4.7) in Cork; by April 30^th^ the values were 10,277 (76.3) and 1156 (21.3). In fact, the rate for Cork on April 30^th^ had occurred 25 days earlier for Dublin.

**Figure 3.**
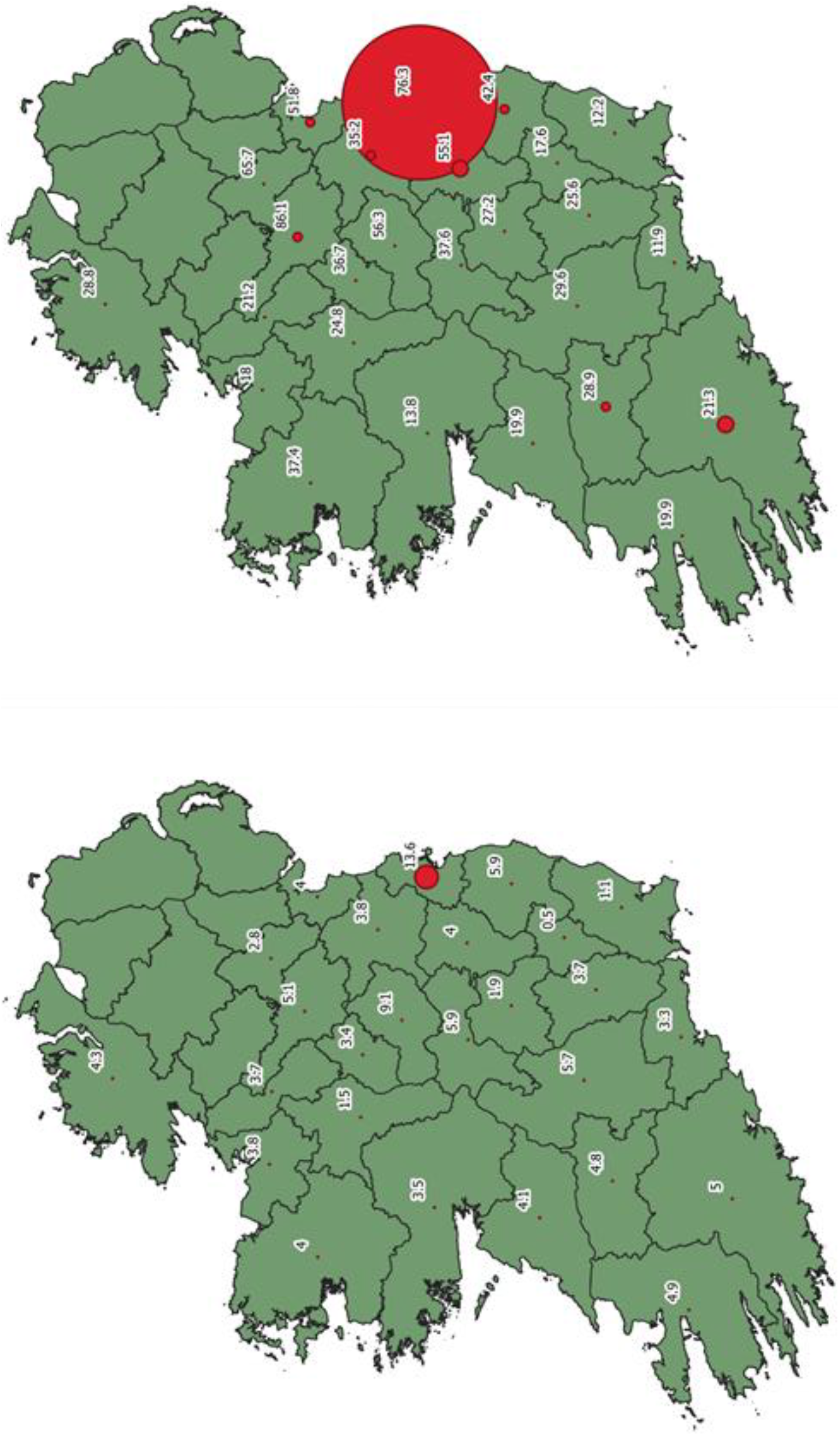
The distribution of cases by county on March 31^st^ and April 30^th^. The size of the proportional symbol represents the number of COVID cases and the label shows the rate per 10,000. For comparison, the number of cases in Dublin on April 30^th^ was 10,277 or > 50% of cases nationally.

Figure 4 shows the rates of infection for selected counties: Dublin, Cork, Galway, Limerick and Waterford, each of which has a large settlement. Note that the values for Dublin are far higher and grow at a much faster rate than the others; in fact, the timeline of rates for Dublin follow the national cases closely. Figure 5 arranges each of the counties by distance from Dublin (based on the geographic centres). Each column indicates the rate of infection on March 31^st^ and on April 30^th^ relative to the median county values of 4.0 and 28.9 per 10,000, respectively. The arrangement of bars indicates that positive values are all closest to Dublin and negative values are further away (Carlow and Laois are exceptions); this pattern has emerged over the study period. This data indicates that the impact of the pandemic is concentrated around Dublin. However, the high rates of infection in the mostly rural counties of Cavan and Monaghan may be linked to transmission across the border with Northern Ireland, which is following a different strategy.

**Figure 4.**
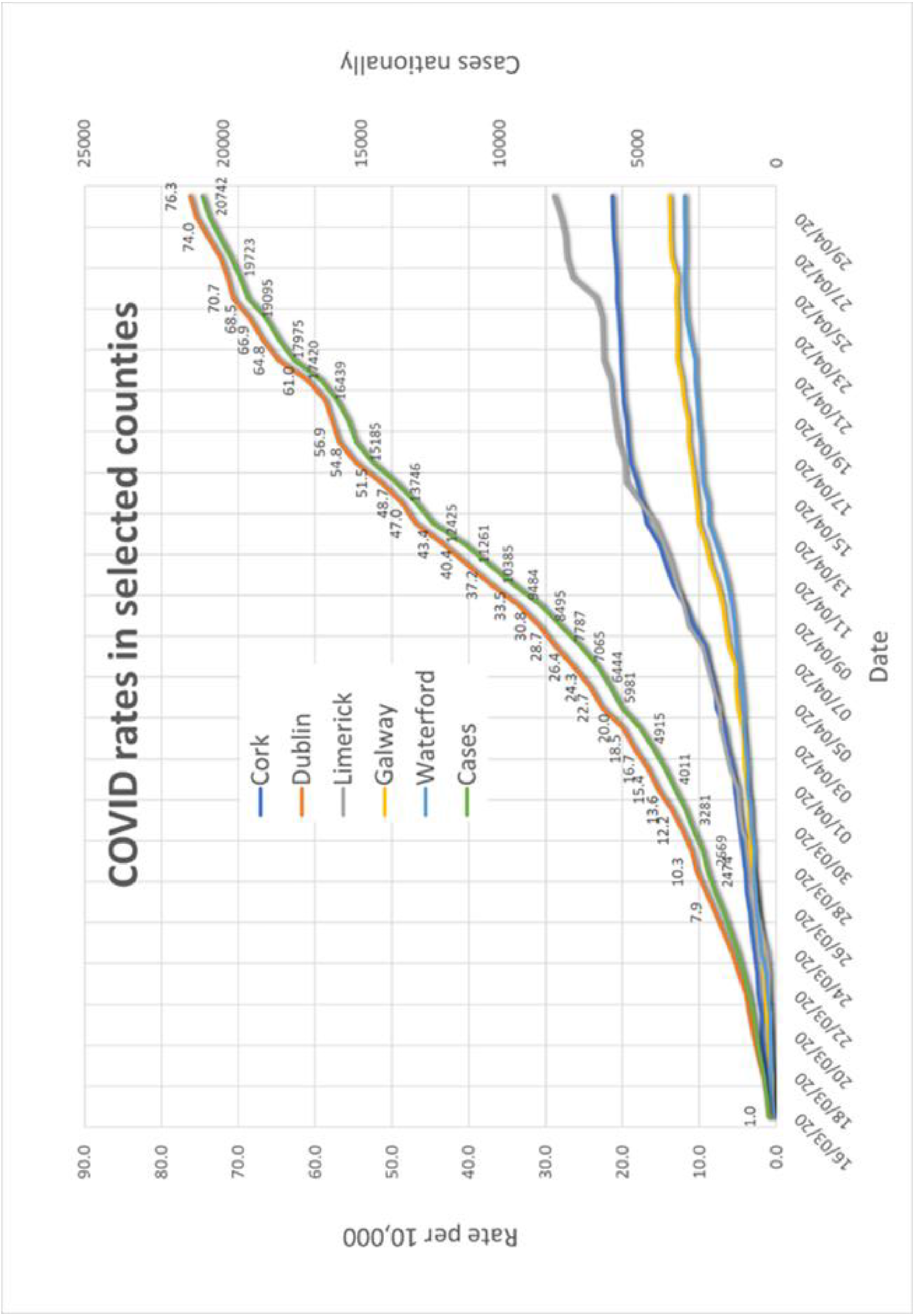
Time series of cases and of rates for selected counties with significant urban populations.

**Figure 5.**
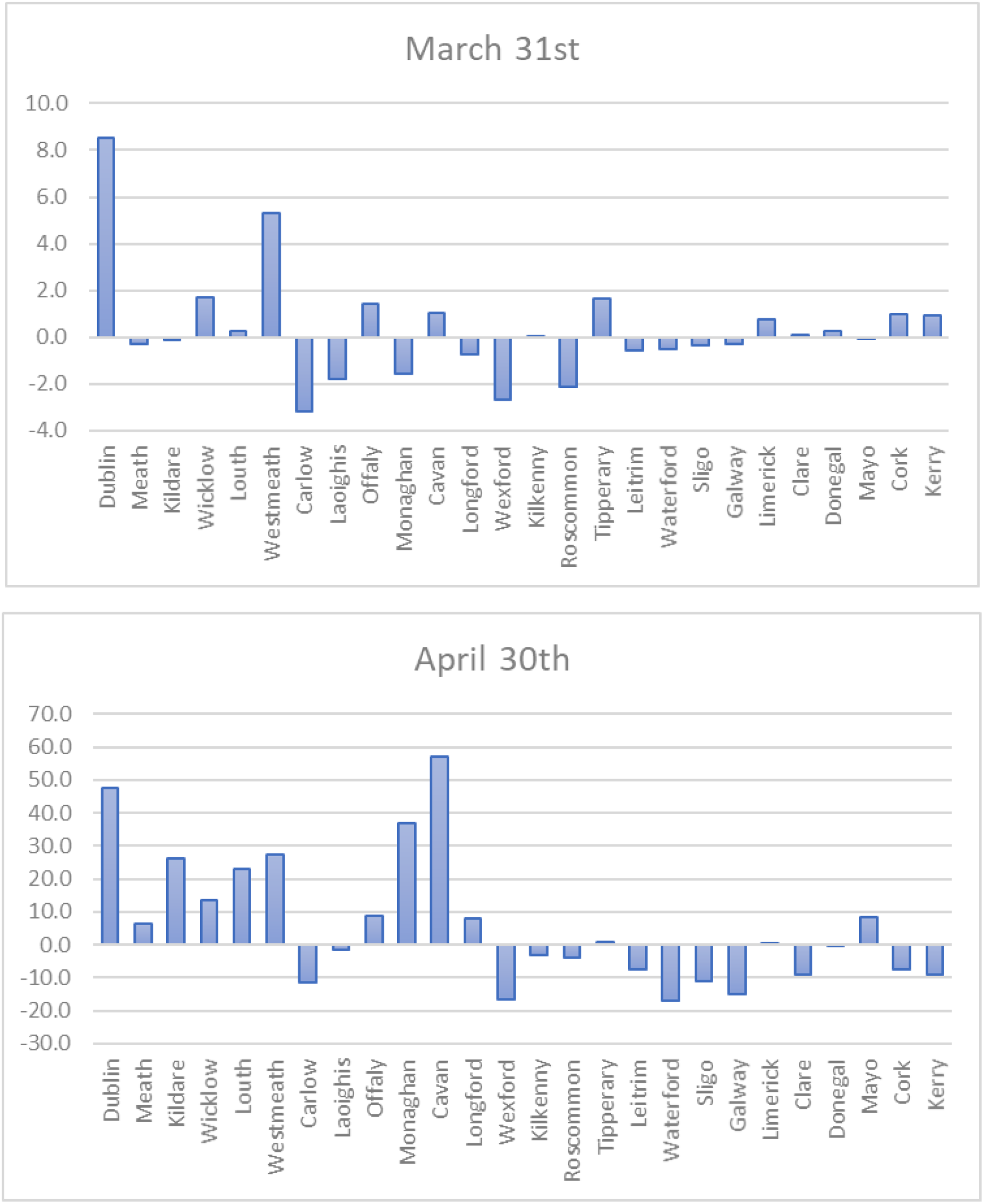
COVID rates for counties compared with the median county rate for March 31^st^ (4.0 per 10,000) and April 30^th^ (28.9 per 10,000), with positive values indicating above average rates. The counties are arranged by distance from Dublin, using the geographic centres of each county.

## Discussion

Understanding how outbreaks of infectious diseases spreads within our communities is important as it helps to identify those areas where primary and secondary prevention measures might best be focussed^6^. The containment phase of the public health response focuses on identifying cases and conducting contact tracing. This process classifies transmission into three categories:

- Community transmission, evidenced by the inability to relate confirmed cases through chains of transmission for a large number of cases, or by increasing positive tests through routine screening of sentinel samples.
- Local transmission indicates locations where the source of infection is within the reporting location.
- Imported cases only indicate locations where all cases have been acquired outside the location of reporting.

The reported data (Table 3) shows that proportion of cases associated with travel have fallen considerably and by April 30^th^ represent just 3% of cases. At the same time the proportion of community transmission has increased.

**Table 3.** How COVID is spreading based on contact tracing at four dates during the study period.

| Date | Community transmission | Local transmission<br>Close contact with confirmed case | Imported cases<br>Travel Abroad | Cases |
| --- | --- | --- | --- | --- |
| March 21<sup>st</sup> | 43 | 24 | 33 | 841 |
| March 31<sup>st</sup> | 59 | 23 | 18 | 3281 |
| April 15<sup>th</sup> | 63 | 32 | 5 | 13012 |
| April 30<sup>th</sup> | 63 | 34 | 3 | 20742 |

An analysis of 308 cases associated with travel that was reported for March 27^th^ indicated that 28% were from the UK, followed by Italy (19%), Austria (17%), Spain (12%) and France (8%)^7^. Almost all travel into Ireland from Europe comes via airline transport and the great majority of passengers pass through Dublin Airport. At the start of 2020 the evidence suggests that the geographic centre of the virus was based in northern Italy and the first cases reported in Ireland were associated with travel to this part of Europe. The spread of disease by global transportation networks has been a topic of study generally^8^. Detailed flight data is not available for the study period but equivalent data for 2019 indicates the distinct nature of travel at this time of the year. For example, in February 2019, just 1% of passengers travelled to and from Austria, yet 17% of the COVID cases in late March originated from here. Many of the trips at this time of year were to ski holiday destinations in the Alps, in places where COVID19 was already in the general population.

The difficulty in containing the pandemic, once introduced into a population, can be illustrated by Figure 6, which shows the home area of those who work in the Dublin Airport census area. The latter includes the Airport (pilots, stewards, security, cleaners) and associated facilities (offices, warehouses, pubs, etc.). These data are from the 2016 Census and show the commuting pattern for a major workplace; note that most workers travel from residences in and around Dublin but there are commuters located along the coasts and in towns and communities across the country, linked by the transportation network. Given that the first cases were ‘imported’ from outside Ireland and passed through the airport^9^, the potential for spread from this location is high.

**Figure 6.**
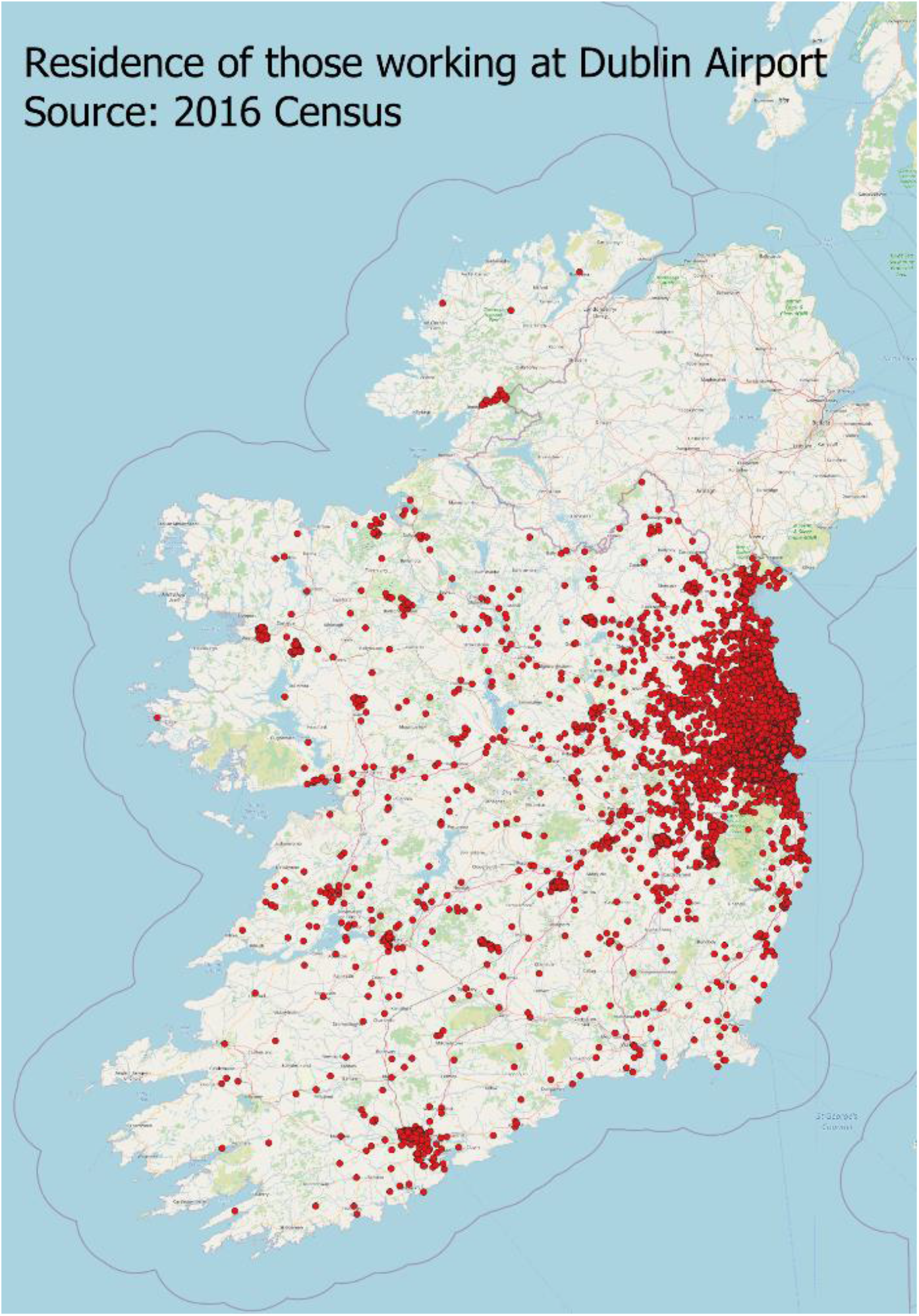
The home areas of those that work in the Airport Electoral District in north Dublin.

On March 9^th^ a decision was made to cancel the annual St Patrick’s Day parade and throughout March, the numbers passing through Dublin Airport fell considerably (Table 4) as fewer people decided to travel.

**Table 4.** Dublin Monthly Passenger Traffic^10^.

| Month | 2019 | 2020 | % Change |
| --- | --- | --- | --- |
| Jan | 2,074,548 | 2,105,782 | 1.51% |
| Feb | 2,009,108 | 2,048,932 | 1.98% |
| Mar | 2,450,960 | 1,044,016 | -57.40% |

The delay phase of the national pandemic strategy officially began in mid-March and has had two parts to this point:

1. March 13^th^ to 29^th^: Cultural institutions, schools, colleges and childcare facilities were closed and mass gatherings were cancelled. People are requested to work remotely if possible.
2. March 28^th^ to May 4^th^: In addition to current restrictions people are urged not to leave their homes apart from those providing an essential service. Others may: shop for food; collect medical prescriptions and medical supplies and attend medical appointments; carry out vital services like caring and; exercise within 2 kilometres of your house. On April 10^th^ the Gardaí were given power to enforce COVID-19 restrictions.

We can assess the effectiveness of these measures, which were designed to slow the rate of transmission, by examining the available on mobility. One source of information is the GPS enabled mobile phone data that is available from Google and Apple. Figure 7 shows the Apple mobility data for Ireland during the study period categorised into driving, transit (public transport) and walking. The scores are presented as an index where 100 represents a benchmark value. The evidence from this source indicates the effectiveness of the policies in greatly reducing the circulation of people.

**Figure 7.**
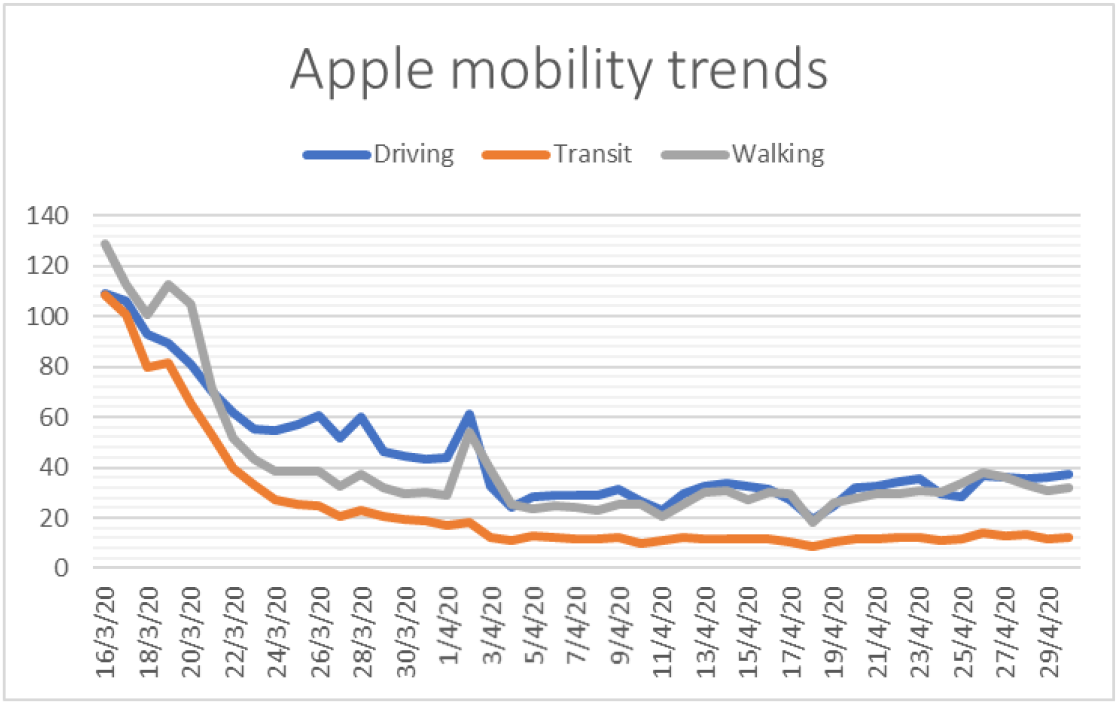
Movements of people as indicated by mobile phone location^11^.

Figure 8 uses traffic data from the National Roads Authority^12^ to examine the impact of the delay strategies on the circulation of people and goods. To make the comparison the morning peak-time traffic for the periods March 2–5 (before the delay restrictions) and April 6–9 (during the delay) were mapped. The patterns show the ‘halo’ effects around each of the cities and large towns and around Dublin in particular, where traffic flows are concentrated; note the high values along the east coast and along major roads into the midlands (compare to Figure 6). The effect of the restrictions has been to reduce the magnitude of traffic by up to two-thirds.

**Figure 8.**
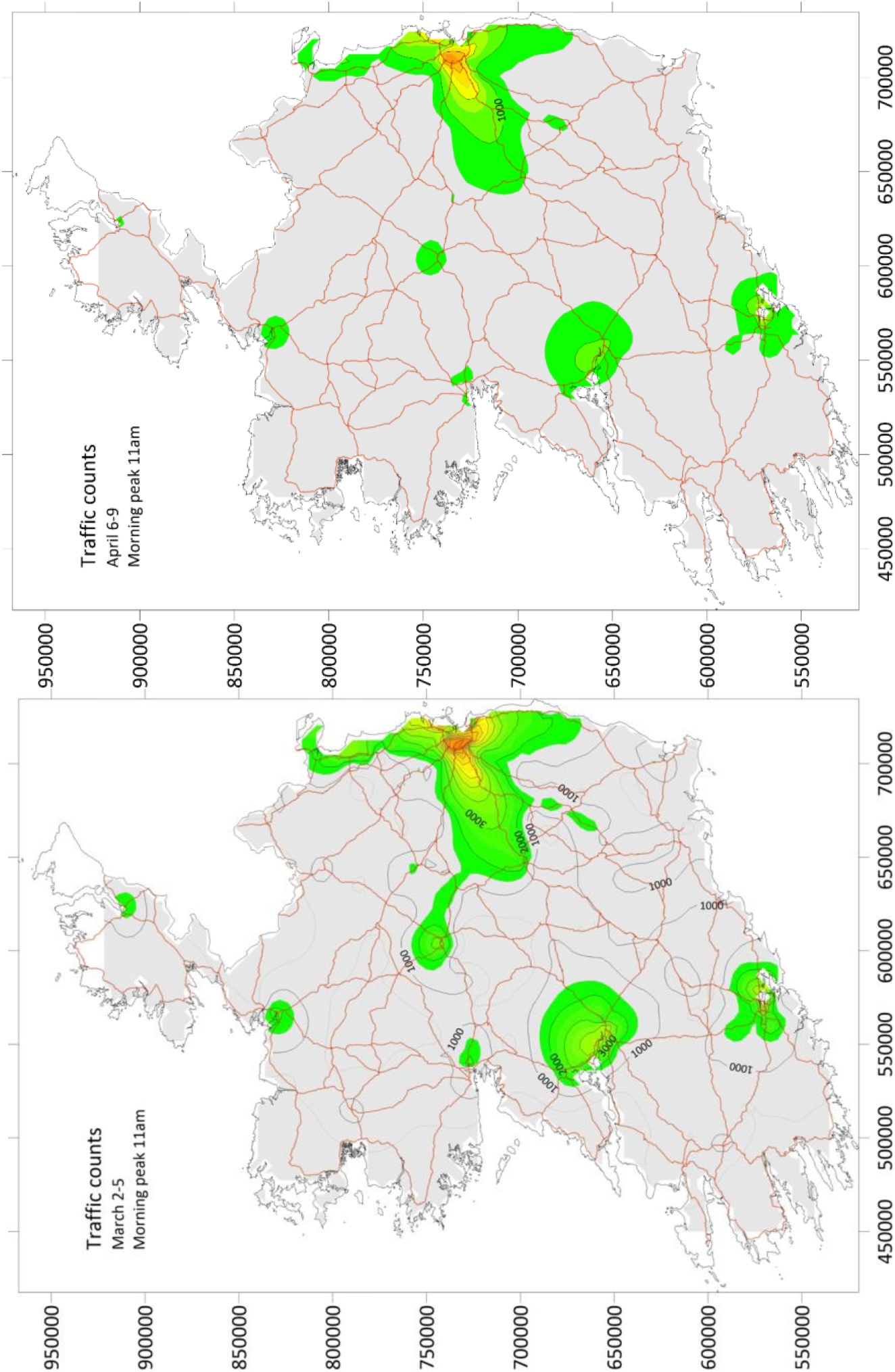
Traffic movements before and during the delay phase of the national COVID strategy. The coloured areas indicate flows of 1000 vehicles or more on the national road system.

The effectiveness of the mobility restrictions (including Garda powers) can be seen in Figure 9. Although the ‘stay-at-home’ restrictions had been in place since March 28^th^, there was concern that travel over the Easter weekend (April 10^th^ – 13^th^) would undermine progress in dealing with the national spread of the pandemic. The figure shows a significant drop-off in travel on the main routes out of Dublin, especially on Friday, April 10^th^, the start of a four-day holiday period.

**Figure 9.**
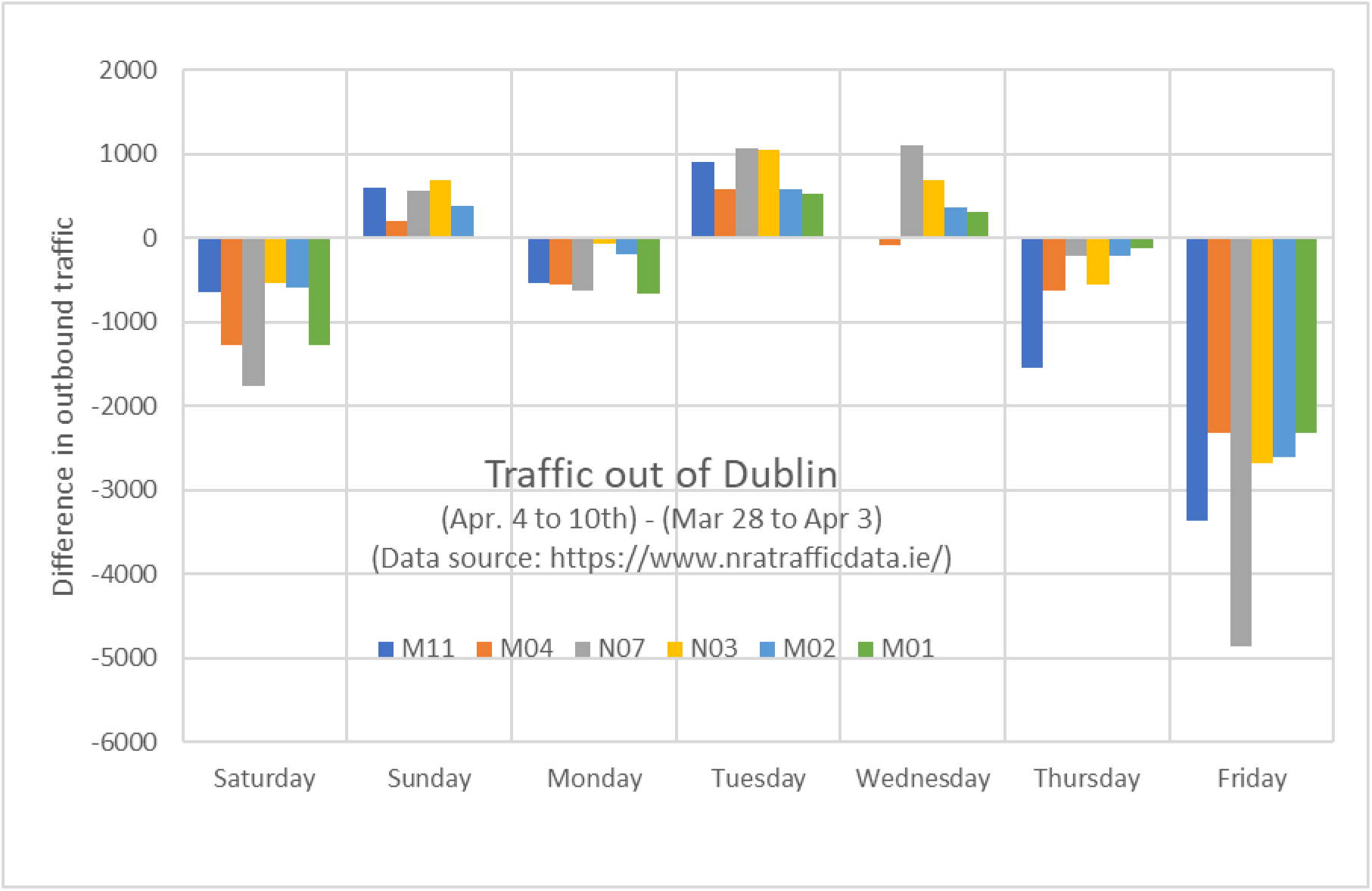
Outbound traffic on the motorways from Dublin over Easter.

It is difficult to gauge the impact of each of the delay strategies on the number of COVID cases simply because the testing system changed over the period as resources were redirected at different points. Based on the reported data, it would appear that the pandemic has become concentrated in clusters that are associated with particular occupations (medical and support staff) and places (hospitals, nursing and community homes); these outbreaks are concentrated in the Eastern (E) region, see Table 5.

**Table 5.**
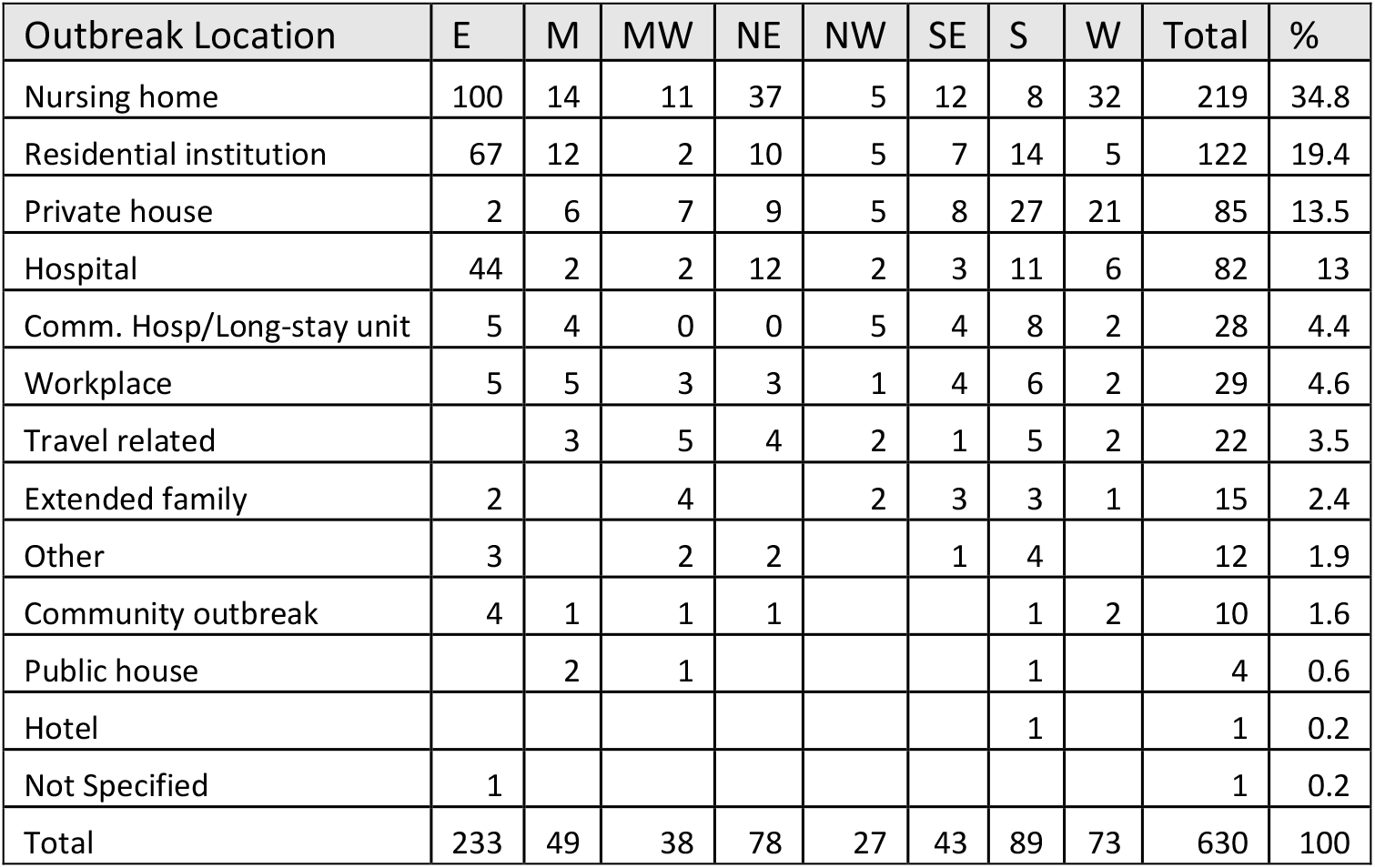
A summary table of outbreaks by location and HSE area for April 30^th^.

Over the study period the number of clusters increased quickly and the average number of cases associated with each cluster increased. Figure 10 shows the time line of clusters and of average number of cases per cluster for the study period. On April 15^th^ these clusters accounted for 20% of reported cases but by April 30^th^ this percent was 33%. Many of these clusters correspond to nursing homes where there is an especially vulnerable population. As of March 31^st^ deaths in nursing homes account for 60% of the national total. To put this in context there are 578 nursing homes in Ireland, 460 of which are either private or voluntary and house and most of which have less than 100 residents. There are also clusters in some work environments such as meat processing plants, which have continued to operate to supply the food chain.

**Figure 10.**
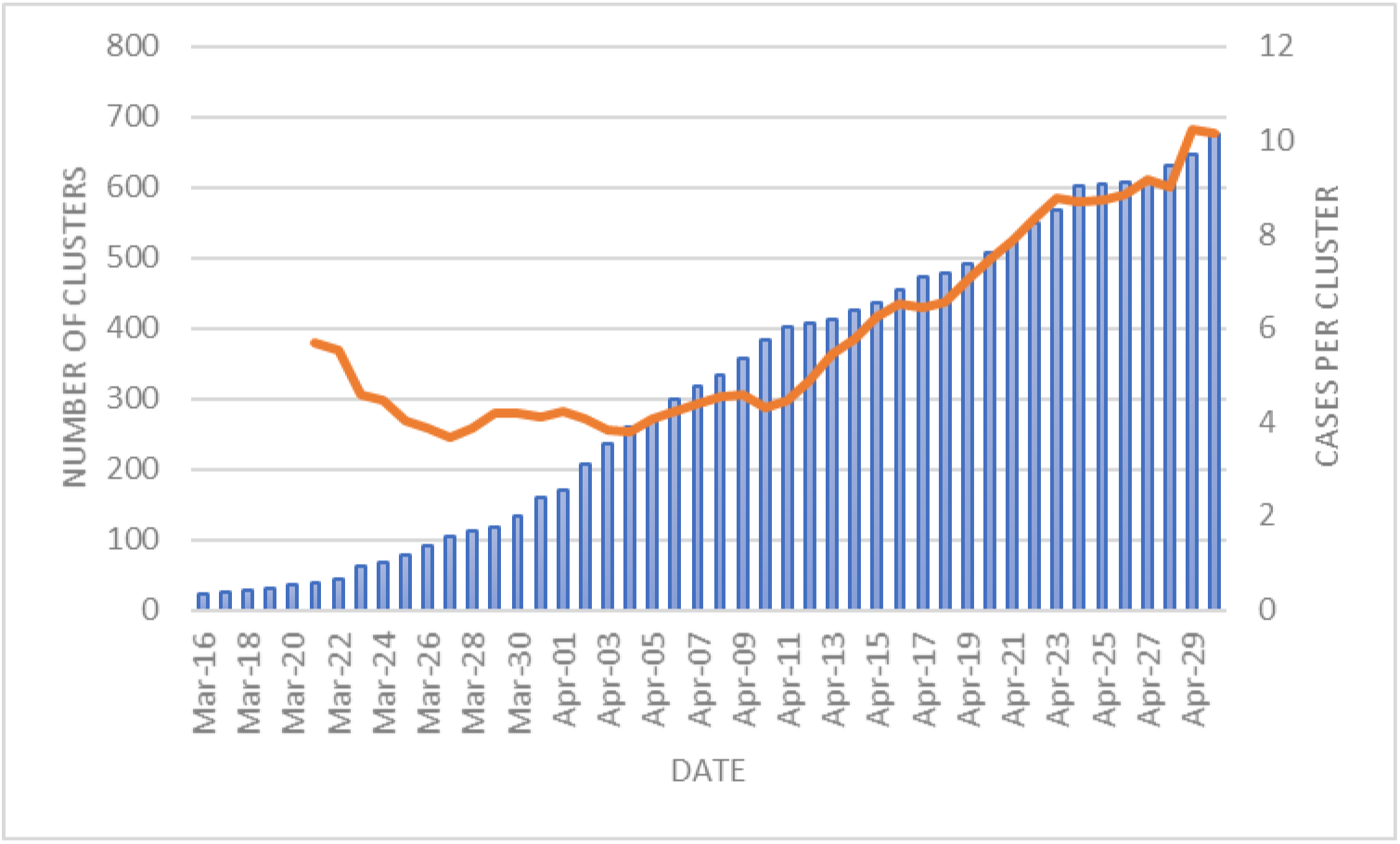
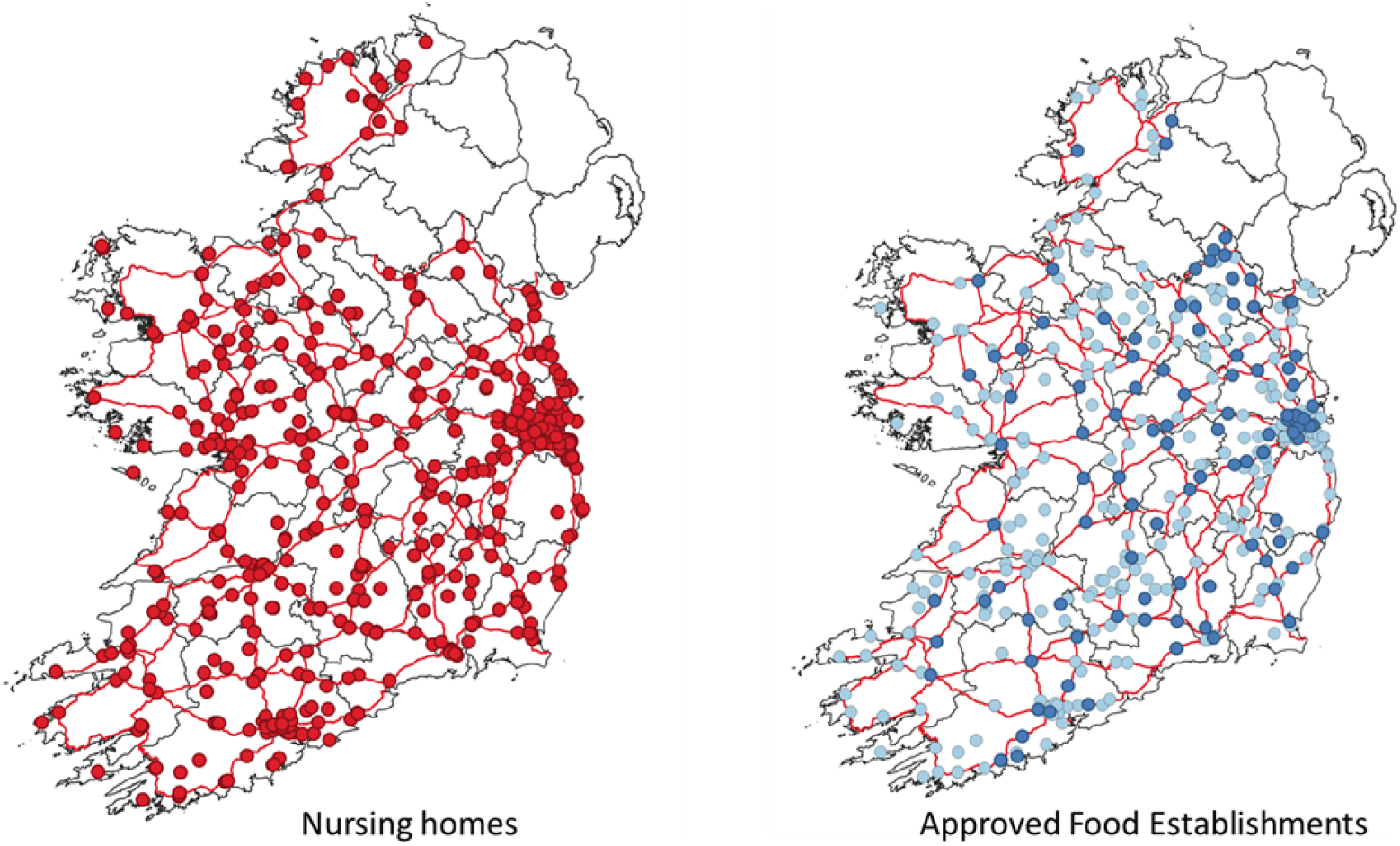
The number of clusters (bars) and the average number of cases per cluster (line) over the study period. The locations of nursing homes and of establishments handling products of animal origin for which there are regulated hygiene conditions. The latter includes meat processing plants.

## Conclusions

Ideally, the public health response to COVID-19 should be linked to examples of good practice elsewhere and informed by local data that is consistent and sufficiently precise. The absence of a coordinated response at a supranational level has meant that each country has followed its own course with different testing, tracing and isolation regimes. This makes it extremely difficult to compare national responses to see which actions are effective at different stages in the spread. This is especially the case in the Europe Union where transport networks and population mobility permit widespread diffusion rapidly. At a minimum, it suggests that there should be a pan-European approach to data gathering and reporting that provide comparable data.

Figure 11 compares the accumulated number of reported COVID-19 deaths for a number of countries in Europe^13^. To allow comparison to with Ireland, the deaths have been standardised to Ireland’s population (4.94 millions) and expressed in terms of deviation from Ireland’s reported deaths; as of 6^th^ May the number of deaths in Ireland were 1375. While Portugal, Germany, Denmark and Greece have a lower number than might be expected, France, UK, Italy, Spain and Belgium have higher values. The reasons for the different outcomes are currently being debated.

**Figure 11.**
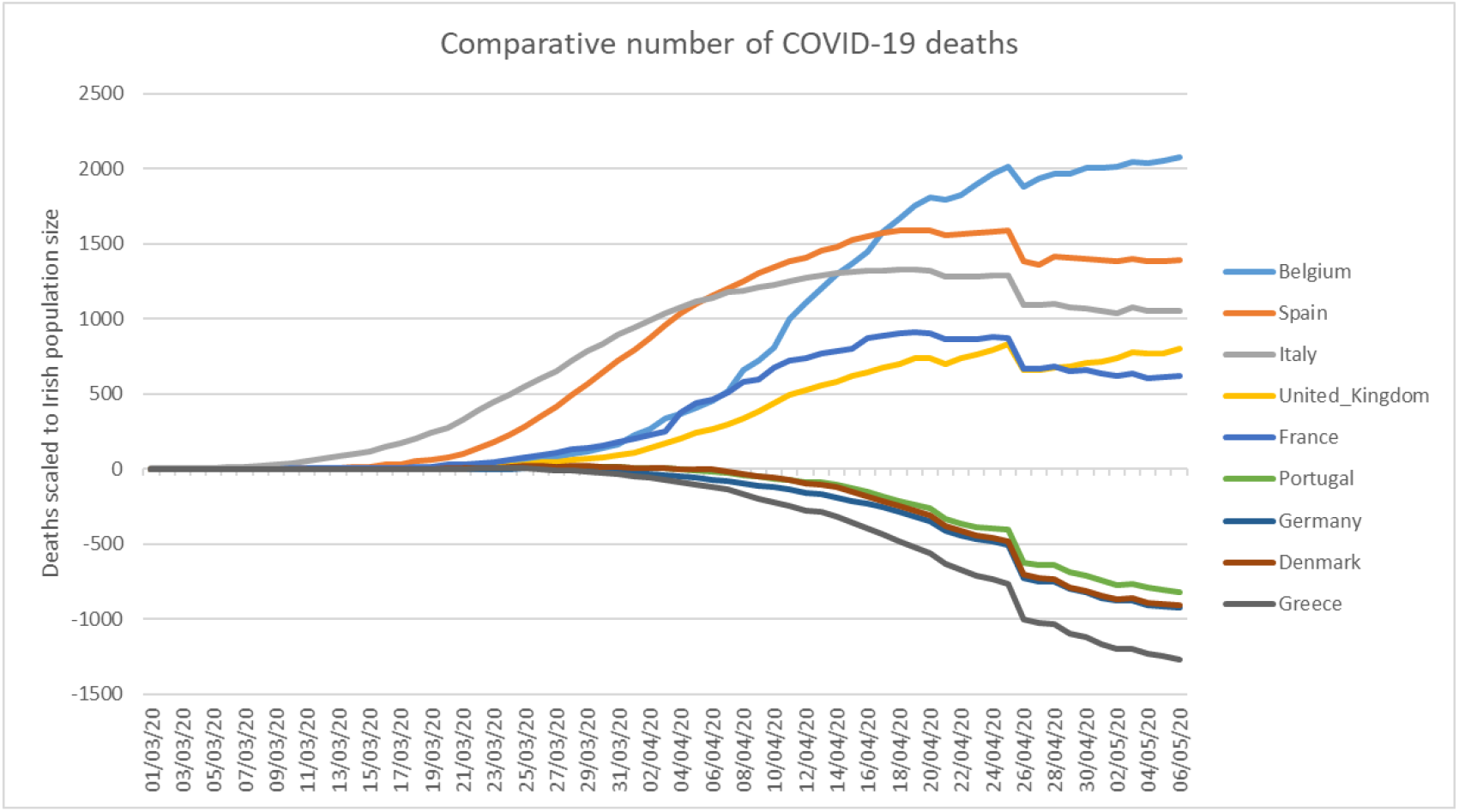
Accumulated COVID 19 deaths across Europe compared to that in Ireland.

To this point, the data for Ireland indicate that:

- Pandemic is concentrated in the east and especially in Dublin and the neighbouring counties. The Dublin focus of the pandemic has become more pronounced over the study period.
- The delay measures that have restricted the mobility of the population have been successful, as measured by the movement of people and traffic. By inference, they have also manged to slow the geographic spread outside of the Dublin area.
- Pandemic has become more concentrated in clusters associated with specific communities (e.g. nursing homes) and professions (healthcare staff).

The caveat in making the above statements is that we are relying on case data primarily to evaluate spread. It is certainly the case that the severe cases are being identified but the rates of infection in the general population are mostly unknown. However, it sems clear that the cases are concentrated in Dublin and adjacent counties and that transmission is now predominantly occurring at a community level. In this respect, the opportunities for spread are likely to be greatest in densely occupied areas characterised by high building and population densities.

The community transmission data globally indicates that the spread of COVID-19 in cities is regulated by higher densities and socio-economic class. The unequal health outcomes for urban communities is a well-established phenomenon but the person-to-person spread of COVID will add to existing place-based health burdens that include poor air quality, impoverished indoor/outdoor environments and embedded social deprivation. Figure 12 shows the population density of Dublin based on the 2016 Census superimposed on the Deprivation Index (DI); the value of the DI indicates socioeconomic status with negative (positive) values corresponding to relative poverty (wealth). Dublin’s city centre and some outlying areas have both high density and high negative DI scores. Moreover, many of those who work in at-risk professions (doctors, nurses, porters, etc.) may live in proximity to their workplace; both the Mater and St. James’ hospital complexes in the city centre are located in high density, deprived parts of the city.

**Figure 12.**
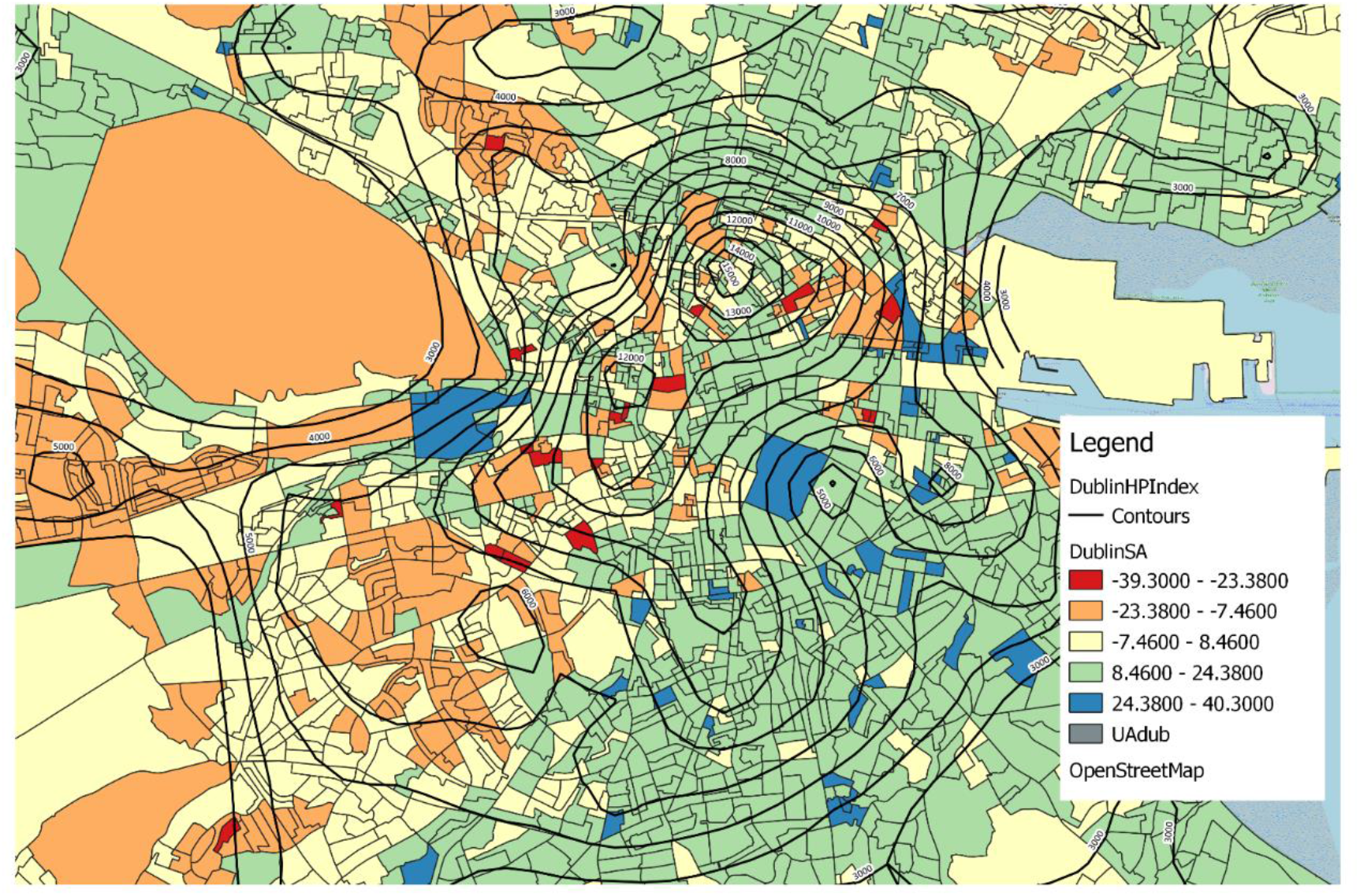
Residential population density (persons per sq.km) in Dublin superimposed over the deprivation index of census areas.

## Next Steps

The nature of the COVID19 pandemic, which is spread by airborne transmission from person to person, affects the organization of a society and economy at a fundamental level. As Ireland emerges from the delay and mitigation phases of this crisis, it must plan for a future in which COVID-19 may be omnipresent and its management requires vigilance. Given its ramifications, the study of the disease cannot be decoupled from its societal context and understanding and managing its spread requires inter-disciplinary approaches. In this short report we have illustrated the potential for collaboration between medical science and social sciences using a range of public datasets to understand the spread of the virus. We hope that this work can provide support for a broad-based approach to short- to long-term planning to create a more resilient society.

### Short term

Internationally, the evidence indicates that as restrictions on population movement and gathering are lifted, there is a strong potential for new COVID outbreaks. To cope with this requires a rapid response that can identify cases quickly and limit community transfer. This requires a testing regime that can detect probable cases rapidly and locate these geographically. More generally, given that many of those infected can be asymptomatic there needs to be random testing to get a measure of the rate of infection in the general population. There is a role for the IT sector (such as mobile phone tracker software) to help in this respect if used in conjunction with public health

### Medium term

In the short to medium term, it should be possible to map COVID risk at a detailed geographic level to identify those circumstances likely to enhance the spread of the virus. These would include densely occupied parts of cities and town, communities that have strong links to occupations at risk (such as those working in health-care facilities), and places where underlying health issues and age makes the population vulnerable. In this respect, information on COVID cases needs to be available at useful geographic scales. For a great many purposes, the Census provides the context for evaluating risk and the reporting level should be commensurate with the Small Areas or at least Electoral District level. The Census data are used by a host of government agencies and local authorities to plan service provision and to identify areas of deprivation.

### Long term

The COVID19 pandemic has revealed the fragility of much of society’s hard and soft infrastructure. The biggest challenge facing use is to develop more resilient services (healthcare, education, transport, etc.) and places of work, home and leisure. For example, high-density cities where significant numbers occupy apartment buildings places great emphasis on the design of neighbourhoods to ensure access to open spaces on an equitable basis. Similarly, the potential for much of the economy and public services to go ‘online’ is predicated on access to IT and overcoming the digital divide. COVID has resulted in some benefits that should not be discounted in long term plans, these include the dramatic improvement in air quality in cities and the value of neighbourhoods as refuges.

## Data Availability

All of the data used in the paper are publicly available from government websites.

## Footnotes

1 https://www.gov.ie/en/news/7e0924-latest-updates-on-covid-19-coronavirus/

2 https://www.hse.ie/eng/services/news/newsfeatures/covid19-updates/

3 https://www.cso.ie/en/index.html

4 https://www.nratrafficdata.ie/

5 https://www.gov.ie/en/publication/20f2e0-updates-on-covid-19-coronavirus-since-january-2020/

6 For example: Beam A, Goede D, Fox A, et al. A Porcine Epidemic Diarrhea Virus Outbreak in One Geographic Region of the United States: Descriptive Epidemiology and Investigation of the Possibility of Airborne Virus Spread. PLoS One 2015;10(12):e0144818. doi: 10.1371/journal.pone.0144818; 2. Adamson WE, Maddi S, Robertson C, et al. 2009 pandemic influenza A(H1N1) virus in Scotland: geographically variable immunity in Spring 2010, following the winter outbreak. Euro Surveill 2010;15(24); Page AL, Ciglenecki I, Jasmin ER, et al. Geographic distribution and mortality risk factors during the cholera outbreak in a rural region of Haiti, 2010–2011. PLoS Negl Trop Dis 2015;9(3):e0003605. doi:10.1371/journal.pntd.0003605 and; Hicks JT, Lee DH, Duvvuri VR, et al. Agricultural and geographic factors shaped the North American 2015 highly pathogenic avian influenza H5N2 outbreak. PLoS Pathog 2020;16(1):e1007857. doi: 10.1371/journal.ppat.1007857

7 Epidemiology of COVID-19 in Ireland. Report prepared by HPSC on 29/03/2020 for NPHET

8 Tatem, A.J., Rogers, D.J. and Hay, S.I., 2006. Global transport networks and infectious disease spread. Advances in parasitology, 62, pp.293343.

9 https://www.rte.ie/news/2020/0320/1124382-covid-19-ireland-timeline/

10 https://www.gov.ie/en/collection/0e1fd9-dttas-quarterly-aviation-statistics-snapshot/

11 https://www.apple.com/covid19/mobility

12 https://www.nratrafficdata.ie/

13 https://www.worldometers.info/coronavirus/

